# Student learning performance and satisfaction with a flipped classroom in undergraduate dental pharmacology education

**DOI:** 10.1101/2024.06.07.24308600

**Authors:** Shelia Galvin, Margaret Lucitt

## Abstract

**Introduction:** The flipped classroom (FC) model of blended learning has become more feasible with the advancement of digital technology platforms. Monitoring a FC approach in curriculum delivery provides an opportunity to evaluate its impact on student exam outcomes and satisfaction. Presented here is examination performances and learning experiences in undergraduate dental students taking pharmacological course material using a FC approach compared to that of a traditional classroom (TC) model.

**Method:** Ninety- seven students experiencing a FC delivery is compared to 129 students taking a TC approach over 2 academic years. Course lecture topics, scheduling and assessment are consistent across both modes of delivery. At the end of each academic year an anonymous student survey with a closed end question was conducted to gain student feedback regarding course satisfaction.

**Results:** The FC positively improved student examination performances compared to the TC approach with a seven percent increase in the percentage mean exam grade and a 15% increase in the number of students obtaining exam grades greater than 50%. An increase in the proportion of students achieving higher grades overall is seen in the FC versus the TC approach from the frequency distribution of exam results. The students also rated the FC more positively in the feedback satisfaction compared to the TC student cohort.

**Conclusion:** In summary the student exam grades and feedback here indicate the FC having a positive impact on student outcomes and experience compared to the TC approach. These findings provide evidence to dental pharmacological educators that a FC curriculum delivery can lead to an improvement in student performances in this subject area.

## Introduction

The flipped classroom (FC) is a teaching approach that is moving away from the traditional classroom (TC) model of lecture based instruction (1) and is being implemented more across higher education institutions since disruptions occurred during the COVID-19 pandemic (2, 3). The TC approach involves the in class didactic lecture where students are usually presented with new content for the first time (4). The TC learning environment is instructor focused and students are passive listeners with little student participation and active learning (5). This form of delivery reduces student participation in class and has been shown to be less effective compared to more active FC learning environments (6-8).

In a FC, students are introduced to the subject matter before in class time through the use of multimedia resources (9) as well as other reading material. Class time is then used for active learning activities such as discussions, problem-solving, group work, and hands-on exercises (10). The pre-class activities allow students to engage with learning material independently before coming to class and typically involve watching video lectures, reading assigned texts, or completing online module problems and case studies. During the in-classroom phase more time is allowed for instructor student interaction, peer group discussions and the application of knowledge (11). The role of the instructor in the FC shifts from delivering content in lectures to guiding students’ understanding, providing feedback, and facilitating deeper exploration of concepts during class time (12). The FC approach emphasizes active engagement and student-centred learning and is now implemented across many health and medical science curricula (13-16). Students have the opportunity to take more responsibility for their learning by actively participating in discussions, asking questions, and collaborating with peers. The flexibility of the FC model also allows for students to engage with online content in a time suitable to them (17). Technology plays a central role in the FC model, enabling students to access learning materials online and facilitating communication and collaboration both inside and outside the classroom (18).

The landscape of higher education teaching delivery across medical and dental curricula is changing as described above from a TC to a FC approach with the introduction of various digital multimedia platforms available to students. As no defined structure to the FC exists, educators are free to implement models to their area of study and subject content when moving away from the TC approach. It is therefore critical to evaluate the learning effects when such models are implemented. During the evaluation process of the FC model it is necessary to understand its potential benefits to student performance outcomes such as in exam grades but also to understand the students’ perception of the learning experience. In order to address this we compared the FC to TC approach in delivery of pharmacological course content to undergraduate dental students evaluating both students learning performances and course satisfaction. The primary aim of this study was to assess the impact of the introduction of a FC model in replacement of a TC approach to deliver pharmacology content to dental students’ learning experience at the Dublin Dental University Hospital, Trinity College Dublin. Specifically, the assessment of student performance focused on a review of exam results related to pharmacological content to measure the impact of a change from a TC to a FC approach in curriculum delivery. This was followed up by an anonymous student survey taken at the end of a continuous FC or TC teaching strategy to measure student perceptions.

## Methods

### Module organisation and delivery

A flipped classroom (FC) and traditional classroom (TC) model in the delivery of a pharmacology curriculum to undergraduate dental science students at the Dublin Dental University Hospital, Trinity College Dublin is described here. The FC design involves a hybrid asynchronous teaching approach compared to the TC synchronous in class lecture based delivery. The course lecture topics, scheduling and faculty members involved in teaching are consistent across both modes of delivery which run over 2 semesters in year 3 of the curriculum. Students experiencing the FC delivery include academic years beginning September 2021 and 2022 with n=97 students in total. Students experiencing the TC delivery include academic years beginning September 2017 and 2018 with n = 129 students in total. Excluded here is academic years beginning 2019 and 2020 due to COVID -19 pandemic disruptions in the examination evaluation format.

### Teaching methods

All content was presented sequentially across both methods of delivery. The FC approach consisted primarily of voiceover lecture slides recorded using Panopto (Panopto, In. Pittsburgh, PA 15212) video platform with reflective questions embedded in the recording for students to review and answer as the recording progressed in real time. Each recorded lecture topic was on average 40min and was released weekly to the students via the virtual learning platform, Blackboard^®^□Learn, over 2 semesters. Lecture notes, textbook chapter reading material and dental clinical case scenarios with reflective questions accompanied each lecture recording for student consideration and pre-class study activities. Live in-person classes with the course lecturer were scheduled every 3 weeks throughout term for review and reflection of released content. Here, students discussed and reflected on prior released content with discussions focusing on pre-class activities. At the beginning of semester 1, the course lecturer presented an in-person introduction to students describing the FC, its layout, pre-class activities, location of content as well as student expectations and engagement. The traditional in-person classroom model consisted of in-class one hour lecture time slots, delivered using PowerPoint presentations followed by a wrap up question and answer time. In the traditional classroom no pre-class activities were required by students in preparation for class instead lecture notes, textbook chapter reading material were released and referred to following each live lecture for students’ to support study activities post in class time.

### Student learning outcomes

Written final semester assessment data related to the pharmacological course content included an extended matching multiple choice style question and a short answer written question. The assessment format was the same across both methods of content delivery. Anonymised unadjusted percentage examination results related to pharmacological course content questions was averaged for each student with an overall average across each type of delivery method taken for comparison. Exam grade data was tested for normality using the D’Agostino-Pearson normality test taking a p value greater than 0.05, as normally distributed. An unpaired students-t-test was used to measure any significant differences. Individual student exam results were also ranked to see the number of students obtaining results above or below the 50% pass grade. Frequency distribution histogram plots showing percentage frequency of students obtaining specified grades between 0-100 % using a 5 percent incremental grade range for each method of delivery was also compared. Here the minimum, median, maximum as well as 25th and 75th percentile grade values were used for comparison in learning performances.

### Student feedback

Feedback data was collected using Qualtrics software (Provo, UT) at the end of each academic year. Students were given a Qualtrics link to an anonymous feedback survey and time in class was allocated to complete. A trichotomous closed end question scale was presented to students to score student satisfaction with the module as: good, satisfactory, or non-satisfactory. Survey responses were averaged for each score across each method of delivery and compared. Student response number of 78.34% (76/97) for the FC versus 62.79% (81/129) for TC were included. The Research Ethics Committee in the School of Dental Science, Trinity College Dublin reviewed and approved the study design.

### Statistical Analysis

Graphical data and statistical analyses were conducted using Excel and GraphPad Prism (version 10.0,GraphPad Software, San Diego, CA, USA). Data for comparison are presented as mean ±□SD and tested for normality using the D’Agostino-Pearson test. Normally distributed data was further analysed using an unpaired two-tailed *t*-test and significant differences taken as * *p* ≤ 0.05, ** *p* ≤ 0.01, *** *p* ≤ 0.001. Chi-square test was used to test significant differences in the response satisfaction rating when comparing the 2 teaching approaches.

## Results

### Student examination outcomes

Presented here is student examination performances in the subject of pharmacology using a flipped classroom (FC) pedagogy model compared to the traditional classroom (TC). In total ninety-seven students were enrolled in the FC versus one hundred and twenty nine students in TC approach. Student examination performances is shown in Figure 1 with the percentage mean ±□SD exam grade in the FC (59.38 ± 1.62 %) significantly greater than that of the TC (51.70 ± 1.45 %) *(p < 0*.*001)*. The FC also positively improved student pass rate (>50%) performances as shown in Figure 2 where 75% of the students taking a FC approach obtained an exam grade greater than 50% compared to 60% of students in the TC approach.

**Figure 1:**
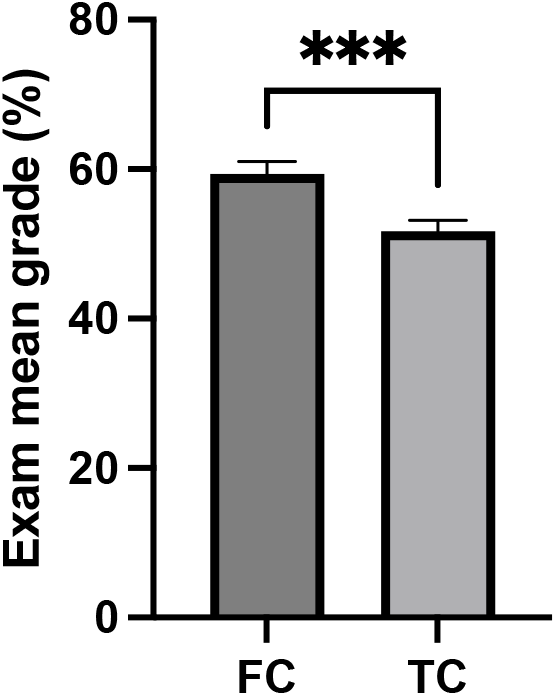
Percentage pharmacological mean exam grades ± SD for students following a flipped classroom (FC) n= 97 versus a traditional classroom (TC) n=129 method of course content delivery. *Statistical analysis of significance between both methods determined using an unpaired students t-test ***p < 0*.*001*.

**Figure 2:**
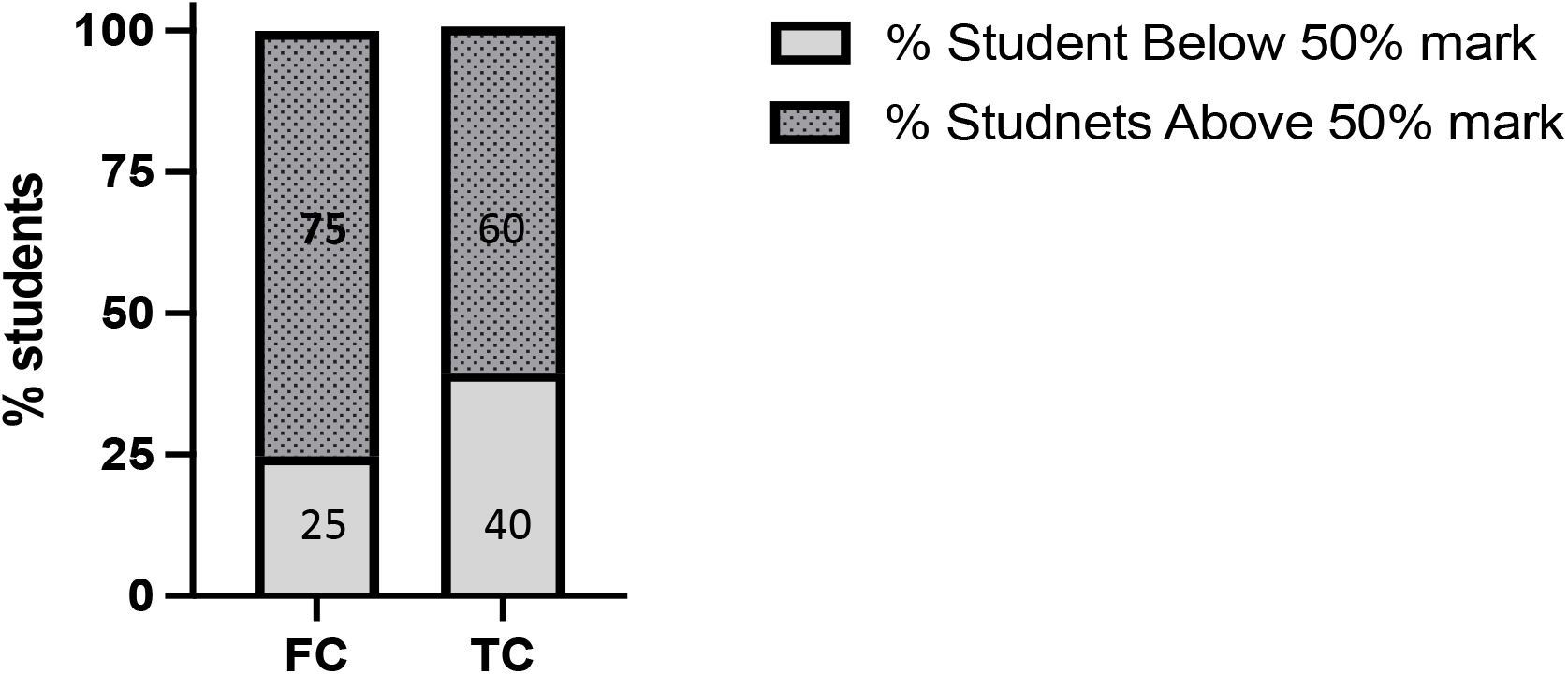
Percentage of students obtaining pharmacological assessment exam grades below 50% and above 50% following a flipped classroom (FC) or traditional classroom (TC) method of course content delivery.

Improvement in exam grades is also shown in Figure 2 with less students in the FC (25% of students) obtaining a grade less than 50% compared to students in the TC (40% of students) approach.

A closer look at the frequency distribution of exam grades for all students is shown in Figure 3. The fitted bell curves for the distribution of results for both FC (Figure 3a) and TC (Figure 3b) display relatively equal normal distributions and this was also calculated with D’Agostino-Pearson normality test (p=0.2548 FC, p=0.4885 TC). Overall distribution of results was improved in the FC approach compared to the TC. This was seen in the most frequent grade in the FC occurring between 70-75% versus 50-55% being the most frequent grade in the TC approach. A minimum exam grade of 5% in seen for the TC which was improved in the FC cohort of students where the minimum exam grade was above 20%. A rightward shift in the fitted bell curve is seen for the FC exam grades (Figure 3a) as evident from the higher percentile grade values (25th percentile grade value of 48.75, median percentile grade value of 60 and 75th percentile grade value of 70) in FC compared to lower percentile grade values (25th percentile grade value of 40, median percentile grade value of 50 and 75th percentile grade value of 60) in the TC distribution (Figure 3b). This rightward shift in the FC curve fitting indicates an increase in the proportion of students achieving higher grades in the FC versus the TC approach overall across all grades. Altogether the FC approach shows an overall improvement in student examination performances compared to the TC approach.

**Figure 3:**
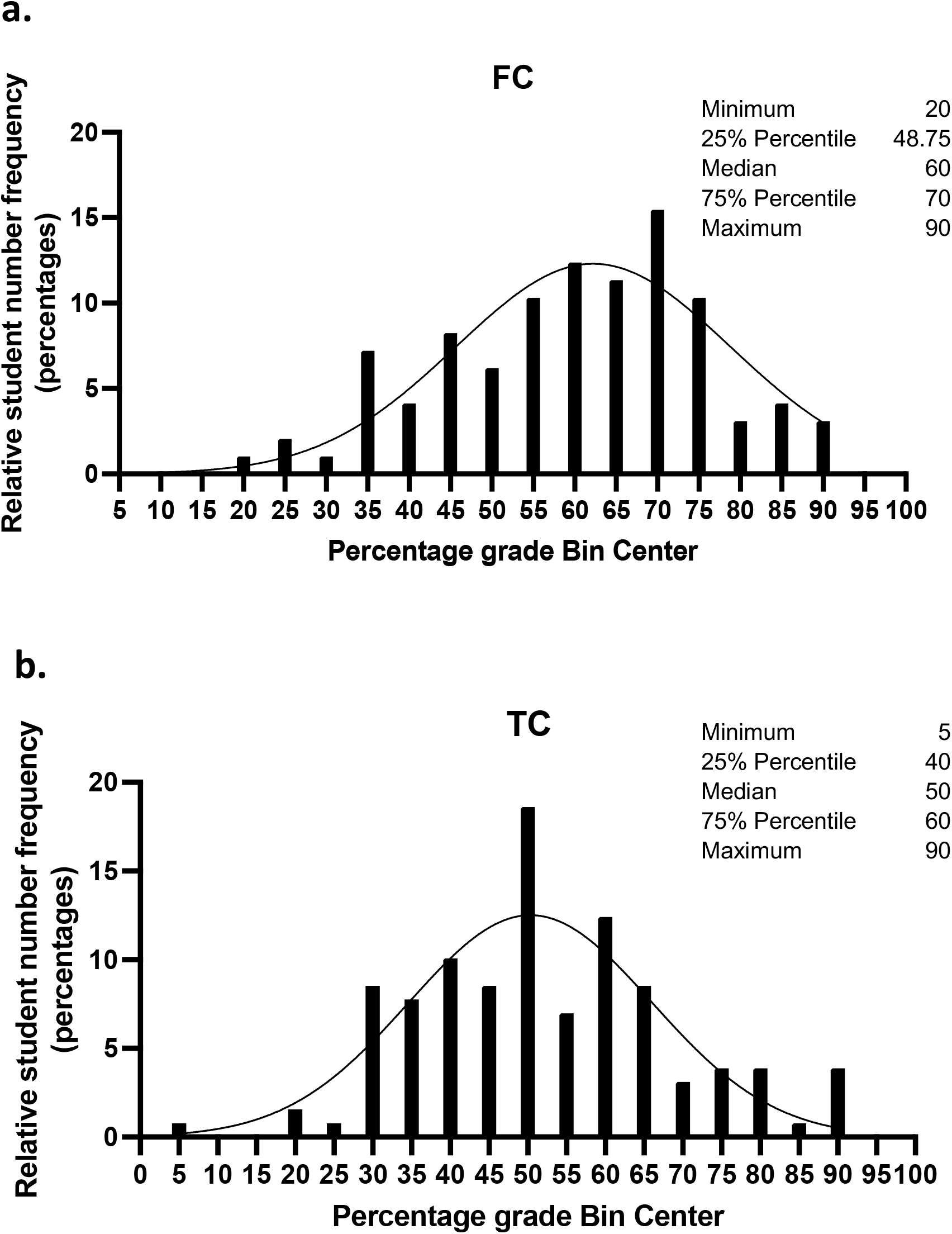
Frequency distribution of pharmacological exam grades obtained by students taking (a) flipped classroom (FC) versus (b) traditional classroom (TC) method of course content delivery. Exam results ranging from 0-100% are plotted with each bar width representing a percentage grade range in increments of 5% versus each bar height representing the frequency of students attaining results for each grade range. Grade values of minimum, mean, maximum, and percentiles (25% and 75%) are displayed on each graph.

#### Student feedback

The student feedback on course content and experience from the anonymous student survey was conducted for academic years beginning September 2017 and 2018 for TC and September 2021 and 2022 for FC shown here in Figure 4. The overall response rate to the student survey was 78.34% (76/97) for the FC versus 62.79% (81/129) for the TC. In general students rated the course content and experience six percent higher in the FC (88% satisfactory or above, raw count) compared to the TC (82% satisfactory or above). This was reflective in the unsatisfactory rating with the TC (17.28%) having a six percent higher score rate compared to the FC (11.82%) approach. This improvement in overall student satisfaction was also seen in the good rating with nine percent higher score rates in the FC (31.5 %) compared to the TC (22.2%) cohort. Despite this a Chi-square test of raw count responses did not show significant difference (p=0.3365) in the response satisfaction ratings when comparing the 2 teaching approaches. Altogether the students had an overall tendency to rate the FC higher compared to the TC student cohort in the satisfactory scaling presented here.

**Figure 4:**
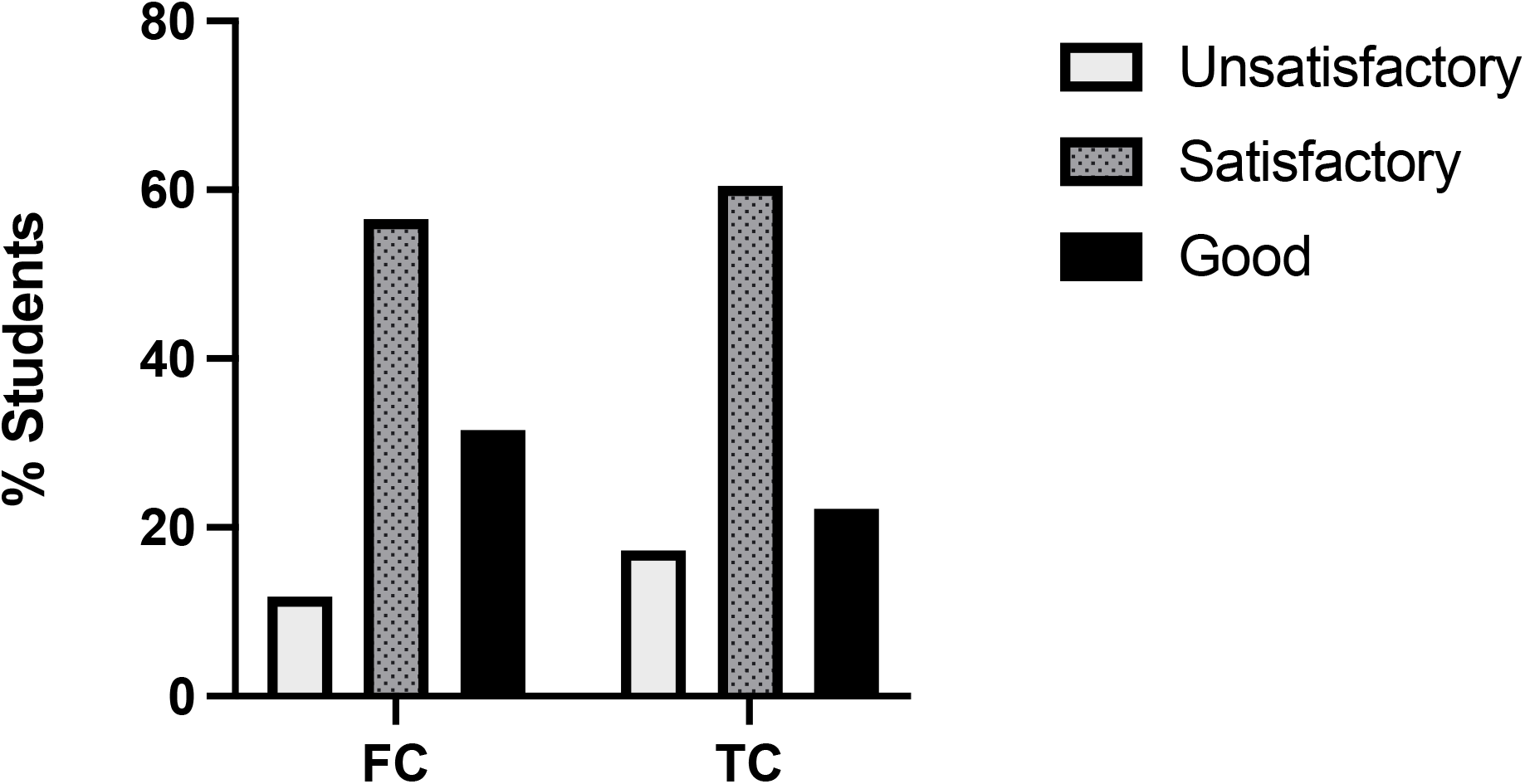
Student feedback based on percentage of student satisfaction ratings from those taking a flipped classroom (FC) versus a traditional classroom (TC) approach. Presented here is percentage of students rating the course as either unsatisfactory, satisfactory, or good.

## Discussion

The idea and model of the FC was first introduced in the 1990s by a Harvard Professor Eric Mazur where he provided material for students to prepare and reflect on before class. This approach allowed for and encouraged deeper cognitive thinking via peer interaction and instructor challenge during in class time (19). Many FC approaches have since been described in the educational literature with a common theme that students have access to some form of course material before in class time (20, 21). The FC teaching approaches today are further facilitated with the advancement and development of digital technology platforms specifically designed for educational institutions (9) and there is now growing evidence on the beneficial effects of the FC model in dental (20, 22) and medical (4, 7) educational programs.

Here we introduced a FC model to our pharmacology undergraduate dental education programme and compared its impact to a TC delivery approach. The mean examination result was significantly better in the FC cohort end of year assessment in pharmacology content compared to the TC cohort (*p < 0*.*001)*. The learning effect of students with the FC approach was better overall with an increase in the frequency of higher grades achieved compared to that of students taking the traditional teaching approach. The FC approach also received more positive feedback from students compared to the traditional instructor-led lecture model.. These findings are consistent with FC approaches applied to other areas of medical and dental educational programs (7, 8, 12, 20).

The overall improved exam results and positive experience from students are likely reflective of the specific aspects of FC teaching, for example, the increased student–instructor interaction and discussion around case-based content introduced in the FC to foster the development of higher-order cognitive skills and application of knowledge (6, 8, 11, 13). The self-directed, independent pre-session time encourages students to take personal responsibility for their own education. This allows students to build a knowledge base before the collaborative classroom sessions empowering students in the learning process. In general students rated the course content and experience higher (satisfactory or above) in the FC compared to the TC despite a Chi-square test of raw count responses not showing a significant difference (p=0.3365) here. This would argue for a mixture of the traditional and flipped classroom models with some students valuing listening to the live in-person lecture (23). A confounding factor here in the satisfaction rating is each student is only experiencing a FC or a TC approach and responses are based on one type of student experience. To truly compare individual students’ experiences and satisfaction a mixed exposure to each teaching approach would provide for a more informative comparison here.

The full effect of the FC educational benefits will only be realised if students engage with the content before class. To help with this, scheduling and timed release of content is important to avoid information overload and overburdening students’ workloads. An ideal approach may also require the use of multiple tools and strategies that accommodate a variety of learning styles while meeting the learning objectives of the curriculum. Careful consideration to factors such as curriculum design and timed delivery, faculty development and the willingness to change teaching approaches, investment in institutional technological infrastructure, staff student ratio in the discussion sessions and student readiness and participation is vital for the successful implementation of the FC model (24, 25). Both student self-directed learning outside the classroom and in-class instructor interactions and discussions is needed to see the full benefits of the enriched environment that can be provided by the FC learning experience. In addition to this designing approaches to teaching and learning should provide for equality, diversity and inclusion in the learning environment that recognises individual differences in the learning process that meets the needs of all students’ effectively. Altogether the positive outcome in learning performances and student experiences the FC model showed in our approach here to the study of pharmacology to undergraduate dental science students suggests the positive impact and utility the FC approach can have.

## Conclusion

The findings here provide evidence to dental pharmacological educators that a FC curriculum delivery can lead to an improvement overall in student learning performances and satisfaction in this subject area. Despite this and the overwhelming literature that support a role for the FC based approach to teaching over the traditional teaching model, it is important that each institution and discipline introducing such changes evaluate its own performances on an ongoing basis.

## Data Availability

All data produced in the present work are contained in the manuscript

## Acknowledgement

The authors would like to gratefully acknowledge dental programme year coordinator Caoimhin Mac Giolla Phadraig for support.

## References

1. Galway LP, Corbett KK, Takaro TK, Tairyan K, Frank E. A novel integration of online and flipped classroom instructional models in public health higher education. BMC Med Educ. 2014;14:181.

2. Blended learning and simulation: the future of dental education in a post-COVID era. Br Dent J. 2022;233(6):490.

3. Namkung J, Goodrich J, Hebert M, Koziol N. Impacts of the COVID-19 pandemic on student learning and opportunity gaps across the 2020–2021 school year: a National Survey of teachers. Front Educ. 2022.

4. Prober CG, Heath C. Lecture halls without lectures--a proposal for medical education. N Engl J Med. 2012;366(18):1657–9.

5. Handelsman J, Ebert-May D, Beichner R, Bruns P, Chang A, DeHaan R, et al. Education, Scientific teaching. Science. 2004;304(5670):521–2.

6. Freeman S, Eddy SL, McDonough M, Smith MK, Okoroafor N, Jordt H, et al. Active learning increases student performance in science, engineering, and mathematics. Proc Natl Acad Sci U S A. 2014;111(23):8410–5.

7. Ji M, Luo Z, Feng D, Xiang Y, Xu J. Short- and Long-Term Influences of Flipped Classroom Teaching in Physiology Course on Medical Students’ Learning Effectiveness. Front Public Health. 2022;10:835810.

8. Wu YY, Liu S, Man Q, Luo FL, Zheng YX, Yang S, et al. Application and Evaluation of the Flipped Classroom Based on Micro-Video Class in Pharmacology Teaching. Front Public Health. 2022;10:838900.

9. Xiao J, Adnan S. Flipped anatomy classroom integrating multimodal digital resources shows positive influence upon students’ experience and learning performance. Anat Sci Educ. 2022;15(6):1086–102.

10. Abeysekera L, Dawson P. Motivation and cognitive load in the flipped classroom: definition, rationale and a call for research.. <i style=““>High Educ Res Dev. 2015.

11. Pierce R, Fox J. Vodcasts and active-learning exercises in a “flipped classroom” model of a renal pharmacotherapy module. Am J Pharm Educ. 2012;76(10):196.

12. Kellesarian SV. Flipping the Dental Anatomy Classroom. Dent J (Basel). 2018;6(3).

13. Joseph MA, Roach EJ, Natarajan J, Karkada S, Cayaban ARR. Flipped classroom improves Omani nursing students performance and satisfaction in anatomy and physiology. BMC Nurs. 2021;20(1):1.

14. Elzainy A, Sadik AE. The impact of flipped classroom: Evaluation of cognitive level and attitude of undergraduate medical students. Ann Anat. 2022;243:151952.

15. Uther P, Van Munster KA, Briggs N, O’Neill S, Kennedy S. Introducing early-phase medical students to clinical paediatrics using simulation and a flipped-classroom. J Paediatr Child Health. 2019;55(9):1107–12.

16. Wang Z, Kohno EY, Fueki K, Ueno T, Inamochi Y, Takada K, et al. Multilevel factor analysis of flipped classroom in dental education: A 3-year randomized controlled trial. PLoS One. 2021;16(9):e0257208.

17. McLaughlin JE, Roth MT, Glatt DM, Gharkholonarehe N, Davidson CA, Griffin LM, et al. The flipped classroom: a course redesign to foster learning and engagement in a health professions school. Acad Med. 2014;89(2):236–43.

18. Nickson CP, Cadogan MD. Free Open Access Medical education (FOAM) for the emergency physician. Emerg Med Australas. 2014;26(1):76–83.

19. Crouch C, Mazur E. Peer Instruction: Ten Years of Experience and Results.. American Journal of Physics 2001.

20. Gianoni-Capenakas S, Lagravere M, Pacheco-Pereira C, Yacyshyn J. Effectiveness and Perceptions of Flipped Learning Model in Dental Education: A Systematic Review. J Dent Educ. 2019;83(8):935–45.

21. Wood DF. Problem based learning. BMJ. 2003;326(7384):328–30.

22. Stenberg E, Milosavljevic A, Götrick B, Lundegren N. Continuing professional development in general dentistry-experiences of an online flipped classroom. Eur J Dent Educ. 2024.

23. Stöhr C, Demazière C, Adawi T. The polarizing effect of the online flipped classroom. Comput Educ. 2020;147.

24. Regmi K, Jones L. A systematic review of the factors - enablers and barriers - affecting e-learning in health sciences education. Bmc Medical Education. 2020;20(1).

25. Hertz B, Clemson HG, Hansen DT, Laurillard D, Murray M, Fernandes L, et al. A pedagogical model for effective online teacher professional development-findings from the Teacher Academy initiative of the European Commission. Eur J Educ. 2022;57(1):142–59.

